# Temporal Dynamics and Interrelations of Cytokines, Neutrophil Proteins, Exudation, and Bacterial Colonization in Epidermal Wound Healing

**DOI:** 10.1101/2023.12.07.23299659

**Authors:** Sigrid Lundgren, Ganna Petruk, Karl Wallblom, José FP Cardoso, Ann-Charlotte Strömdahl, Fredrik Forsberg, Congyu Luo, Bo Nilson, Erik Hartman, Jane Fisher, Manoj Puthia, Karim Saleh, Artur Schmidtchen

**Author notes:** Corresponding author: Artur Schmidtchen, Address: Division of Dermatology, Department of Clinical Sciences, Lund University, Biomedical Center B14, Baravägen 18, SE-22241 Lund, Sweden.

## Abstract

Inflammation is integral to wound healing, yet its dynamics in normally healing epidermal wounds are not fully characterized. To this end, we analyzed longitudinal wound fluid samples collected from epidermal suction blister wounds in healthy volunteers. Cytokine levels peaked on day 5, followed by a decline on day 8. Wound exudation, measured by protein content, increased from day 2, peaking on day 5. The neutrophil-derived proteins myeloperoxidase, elastase, and heparin binding protein (HBP) peaked on day 5, correlating with interleukin (IL)-8, a key neutrophil chemoattractant. With a focus on viable cultivable wound bacteria, aerobic species were cultured from wound swabs and dressings and identified by MALDI-TOF mass spectrometry. The identified bacteria consisted primarily of commensal bacteria, including various staphylococci. Levels of such bacteria rose from day 2, peaking on days 5-8, and aligned with increases in the cytokines tumor necrosis factor (TNF)-α, IL-1β and IL-10 at the peak of inflammation on day 5. The study uncovers the coordinated dynamics of cytokines, neutrophil activity, and major commensal bacteria in epidermal wound healing, and identifies HBP – a marker for neutrophil activation and endothelial leakage – for the first time in normally healing epidermal wounds. The clinical study in which the analyzed samples were collected is registered in clinicaltrials.gov (NCT05378997).

## Introduction

Wound healing is a highly regulated biological process characterized by a series of well-orchestrated events involving inflammation, proliferation, and remodeling, all aimed at restoring tissue integrity and functionality ^1–3^. Inflammation, exudation, and neutrophil responses are all important aspects of wound defense and repair, collectively ensuring that the wound is cleansed of debris and pathogens, creating a conducive environment for tissue repair and regeneration ^3–6^. Central to the early phase of wound healing is the release of key pro-inflammatory cytokines, such as tumor necrosis factor (TNF)-α and interleukin (IL)-6, which promote immune cell recruitment and activation. IL-8, a potent neutrophil chemoattractant, plays a critical role in driving neutrophil migration and function at the wound site. Neutrophils, in turn, can release proteolytic enzymes like neutrophil elastase (NE) and myeloperoxidase (MPO), which contribute to bacterial clearance and tissue remodeling ^5,7,8^.

However, in certain disease states including various types of non-healing wounds, excessive neutrophil activity or prolonged cytokine signaling can lead to tissue damage, highlighting the importance of tightly regulated cytokine and protease dynamics in normal wound healing. Moreover, bacteria can significantly influence these processes by enhancing inflammation, leading to prolonged tissue damage and delayed healing ^9,10^. From a physiological perspective, commensal bacteria can promote skin homeostasis and wound healing^11–15^. However, recent findings challenge the belief that commensal bacteria solely benefit wound healing. While *Staphylococcus epidermidis* colonization can be protective against infections, high-dose applications of commensal bacteria, such as *S. epidermidis*, to abraded skin can delay barrier repair and induce a chronic wound-like inflammatory state^16^. This is consistent with reports that barrier disruption generates an inflammatory environment, which itself promotes pathogen colonization and infection leading to suppression of the protective mechanism of skin commensals ^17^. Together, these results highlight the nuanced role of commensals in modulating wound healing.

Epidermal wounds, specifically affecting the outermost skin layer, present a controlled environment to study these interactions. The suction blister model offers a standardized approach to create uniform wounds ^18–22^, providing valuable insights into the dynamics of inflammation and bacterial colonization during healing. This study was conducted to establish a baseline characterization of inflammatory and bacterial dynamics in epidermal wound healing, providing a foundation for future studies employing the suction blister model or related wound models. We performed a secondary analysis of control samples from a randomized, placebo-controlled clinical trial^23^ (NCT05378997) utilizing the suction blister wound model. We used multiplex Meso Scale analysis, ELISA, MALDI-TOF mass spectrometry and microbiological methods focused on viable, metabolically active commensal bacteria to comprehensively examine cytokine dynamics, neutrophil activity, and bacterial colonization and levels in epidermal wound healing. The study reveals the dynamics of cytokines, neutrophil proteins, and commensal bacteria in epidermal wound healing, identifying HBP, a known marker for neutrophil activation and endothelial leakage, for the first time in normally healing epidermal wounds.

## Materials and methods

### Study design

We analyzed samples collected as part of a randomized, controlled trial designed to test the safety and pharmacokinetics of thrombin-derived C-terminal peptide (TCP)-25 in epidermal suction blister wounds (clinicaltrials.gov identifier NCT05378997). Twenty-four subjects were included in the study with two placebo-treated suction blister wounds each. We only included samples from control placebo-treated wounds in this study. The study design is described in detail in the published study protocol ^23^.

### Formation of epidermal suction blister wounds

Two blister wounds were created on the medial aspect of each thigh using a Model NP-4 (Electronic Diversities, Finksburg, MD) suctioning device ^24^ as described in the published study protocol ^23^. On each thigh, one wound was randomized to receive active drug and the other was randomized to receive control gel without the drug substance. Since drug-treated wounds were not included in this study, we included 48 control wounds from 24 subjects.

### Swab procedure and analysis

The wound was swabbed on day 1 (immediately after blister formation) and on days 2, 3, 5, 8, and 11 using a sterile cotton swab (Selefa, Stockholm, Sweden) and standard methods ^25^, and extracted as described in the supplement. Swabbing is a common technique for collection of bacteria from wounds and skin, yielding bacterial information that is comparable to other methods ^26,27^. Bacterial levels were quantified immediately after extraction as described in the supplement, following standard methods ^25,28^.

### Dressing extraction procedure and analysis

Wounds were covered with Mepilex dressing (Mölnlycke healthcare, Gothenburg, Sweden) in between study visits. Dressings were changed on days 2, 3, 5, 8, and 11. At each dressing change, the dressing was removed from the wound and dressing fluid extracted as described in the supplement. Bacterial levels were quantified immediately after extraction in fresh (never frozen) dressing fluid as described in the supplement. Cytokines (interferon gamma [IFN]-γ, IL-1β, IL-2, IL-4, IL-6, IL-8, IL-10, IL-12p70, IL-13, tumor necrosis factor [TNF]-α), protein content, and neutrophil proteins (neutrophil elastase [NE], myeloperoxidase [MPO], and heparin binding protein [HBP]) were quantified in the stored dressing fluid samples that contained protease inhibitor, as described in the supplement.

### Rationale for the chosen bacterial quantification and identification strategy

The scope of this part of the study was limited to evaluating the cultivable bacterial burden on the wound surface and in dressings, with a particular focus on commensals of normally healing wounds. Therefore, to quantify only the viable cultivable bacteria, quantitative bacterial counts was the method of choice for this study ^28,30^. With a specific aim to identify and discriminate between the major cultivable aerobic bacteria, including *Staphylococcus aureus* and various commensal staphylococci, Matrix Assisted Laser Desorption lonization - Time Of Flight (MALDI-TOF) mass spectrometry (MS) associated with MALDI Biotyper software was used for the rapid and specific identification at the species level ^31,32^.

### Identification of major cultivable bacteria

Swab and dressing fluid samples were streaked on a blood agar plate and six colonies were selected for analysis by MALDI-TOF to identify the major cultivable bacteria, as described in the supplement. Blood agar was selected as the growth medium as is commonly used in clinical microbiology laboratories as it is a rich medium that allows growth of most common clinically relevant pathogens ^28^.

### Wound imaging

A disposable centimeter-scale ruler was placed on the skin near the wounds. Two images were taken of each set of wounds on each leg at a distance of 35 cm with a Canfield Twin Flash camera (Canfield Scientific, Parsippany-Troy Hills, NJ, USA) with a modified version of a Canfield close-up scale ^29^.

### Statistics

Summary data were plotted as median and interquartile range. For correlation between two variables, Spearman correlation coefficients were determined. Although each participant had two control wounds that were included in this study, individual wounds were considered independent biological replicates in all analyses (n = 48 wounds). Due to the non-normal distribution of the data (determined by visual inspection of the data), non-parametric analyses were used throughout. All statistical tests used were two-tailed. Due to the exploratory nature of the study, p-values were not adjusted for multiple comparisons and should be considered hypothesis-generating only. Data were analyzed and plotted using GraphPad Prism 10 software. When a logarithmic axis was used, zero values were replaced with 1 to enable visualization of all data points on the graph but were not replaced in any statistical analyses.

### Ethical considerations

The clinical trial and collection of specimens to the biobank were approved by the Swedish ethical review authority (Etikprövningsmyndigheten application number 2022-00527-01). Written informed consent was received from all subjects prior to participation.

## Results

### Inflammation

To determine how inflammation develops over time in normally healing epidermal wounds, we measured the levels of 10 different cytokines in the dressing fluid on days 2, 3, 5, and 8. Because there was very little exudation on day 11, we did not measure cytokine levels on this day as they were likely to be undetectable. Although the individual cytokines had vastly different levels, ranging from about 1 ng/mL (IL-4) to over 10,000 ng/mL (IL-8) at their peak, all cytokines followed similar dynamics over time (Figure 1). Most measured cytokines remained at approximately the same level from day 2 to 3 (IL-1β, IL-2, IL-8, IL-12p70, IL- 13, TNF-α), while some declined slightly on day 3 relative to day 2 (IL-4, IL-6, IL-10). IFN-γ, in contrast, increased steadily from day 2 to 5. In spite of differences in initial dynamics, all measured cytokines reached their peak level on day 5 and then declined on day 8.

**Figure 1.**
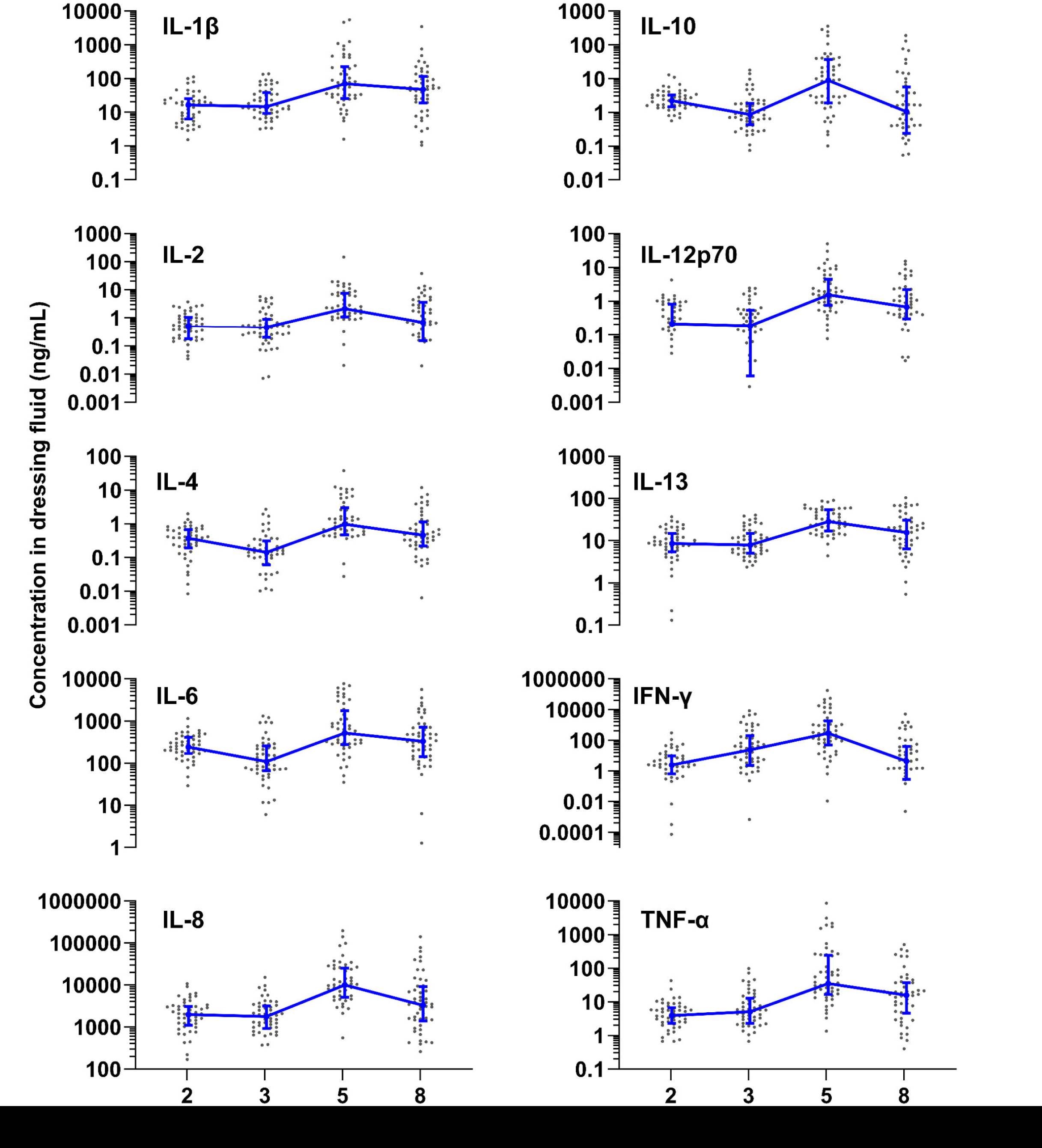
Inflammation over time. Levels of ten different cytokines measured by Meso Scale in dressing fluid over time. Each data point is the wound fluid cytokine concentration in a single wound, while the blue line is the median of all wounds that day (n = 48), and whiskers are the interquartile range.

To confirm that measured cytokine levels are not dependent on the amount of protein content in the dressing fluid, we normalized the levels of each cytokine for the protein content of that same sample (Supplementary Figure 3A). The overall dynamics of the cytokines did not change greatly when normalized for protein content, reaching their peak at day 5. The normalized level of some cytokines (IL-1 β, IL-4, IL-6, IL-12p70, TNF-α) did not decrease on day 8, unlike un-normalized levels.

### Wound exudation

To determine how the level of wound exudation changes over time, we measured protein content in dressing fluid as a proxy for wound fluid accumulation. Because all dressing fluids were extracted with the same volume of extraction buffer, the amount of protein indicates the accumulated amount of wound fluid in the dressing and is not affected by possible evaporation through the Op-Site film covering the dressing. The results indicated that the level of exudate was moderate on days 2 and 3 and then peaked on day 5 before dropping on day 8 (Figure 2A). On day 11, dressings were very dry and had near-zero levels of protein content, indicating very low exudation on this day.

**Figure 2.**
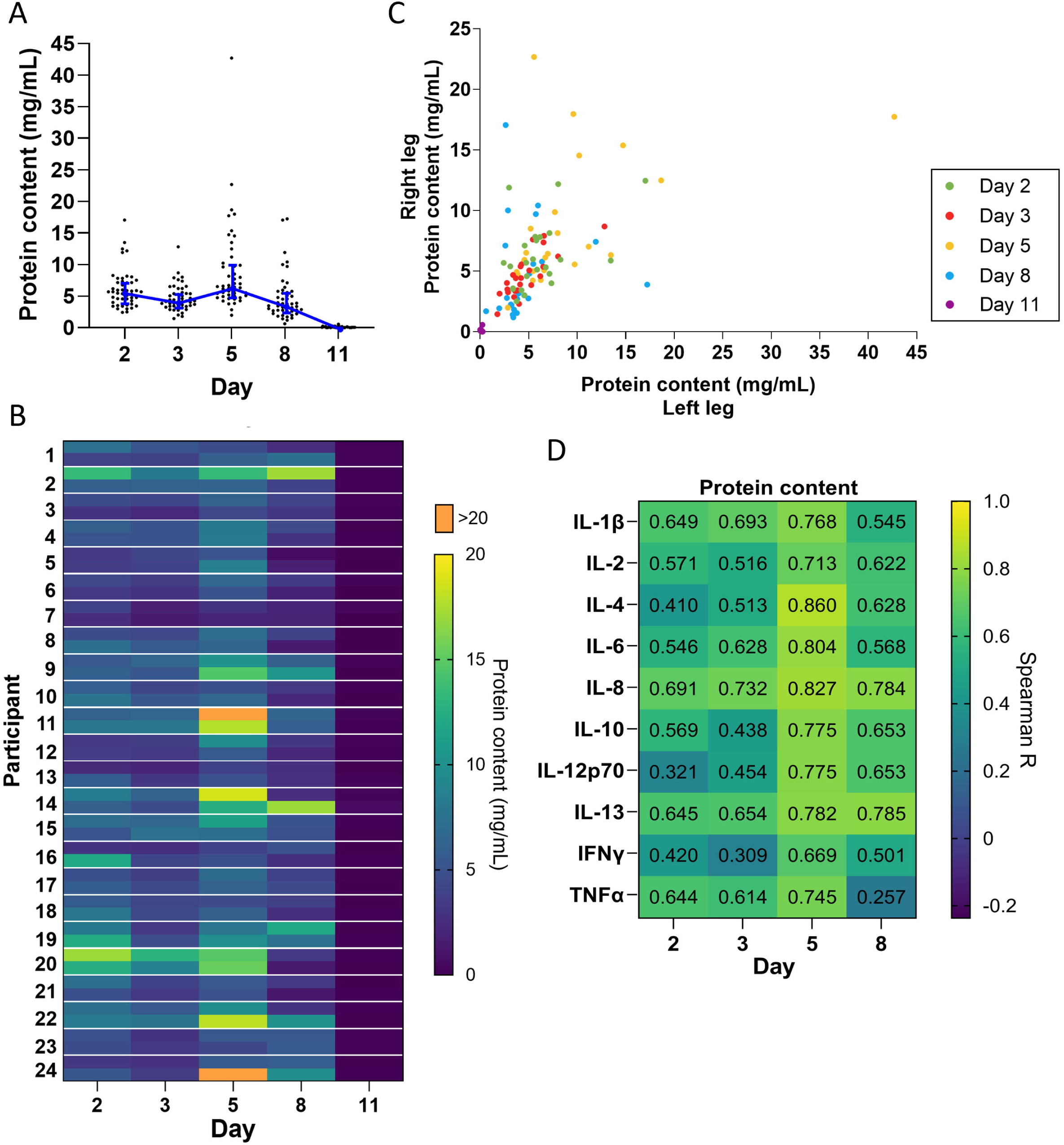
Exudation over time. A) Protein concentration quantified in dressing fluid from each wound at each time point. Each data point is protein concentration in a single wound, while the blue line is the median of all wounds that day (n = 48), and whiskers are the interquartile range. B) Heat map of protein concentration quantified in dressing fluid samples from each wound. The left and right wound are each presented as a single cell on the top and bottom of each row, respectively. C) Scatterplot comparing protein concentration in dressing fluids from wounds on the left leg vs the right leg in each participant. D) Spearman correlation coefficients comparing levels of each cytokine with protein content in that wound (n = 48)

When plotted as a heatmap, protein content showed considerable individual variation, and appeared to be similar between left and right wounds (Figure 2B). To confirm this, we plotted protein levels in the left and right wound as a scatterplot (Figure 2C) and determined the Spearman correlation coefficient on each day and overall (Table 1). The data indicated a moderate to high correlation between left and right wounds on all days (R = 0.356 – 0.803), except for day 11 which had many zero values. The data also indicated a strong correlation overall when all days were included (R = 0.791), indicating that there is some variation in exudation between left and right wounds in each individual as the wound heals, but that overall exudation levels do not differ greatly between wounds on the left and right leg.

**Table 1.** Correlation coefficients for protein content in left vs right leg.

| Day | Spearman R | P value | 95% confidence interval |
| --- | --- | --- | --- |
| Day 2 | 0.409 | 0.0474 | 0.00630 to 0.704 |
| Day 3 | 0.803 | <0.0001 | 0.582 to 0.913 |
| Day 5 | 0.638 | 0.00100 | 0.305 to 0.832 |
| Day 8 | 0.356 | 0.0880 | -0.0683 to 0.671 |
| Day 11 | -0.029 | 0.894 | -0.437 to 0.390 |
| Overall | 0.791 | <0.0001 | 0.710 to 0.851 |

Inflammation is known to induce capillary leakage and subsequent exudation. We determined the correlation coefficient between the level of each cytokine and protein content of the dressing fluid. We found that there was a moderate to strong correlation between cytokine levels and protein content on all days (R = 0.257 to 0.860), particularly on day 5 (R = 0.669 to 0.860) (Figure 2D).

### Neutrophils

Neutrophils play a key role in wound healing and bacterial control. Therefore, we measured the levels of three different neutrophil proteins (MPO, NE and HBP) on days 2, 3, 5, and 8 (Figure 3A). Due to the small amount of exudation on day 11, proteins were likely to be undetectable, so we did not measure neutrophil protein levels on this day. MPO and NE remained at approximately the same level from day 2 to 3, peaking on day 5, and then dropping sharply on day 8. In contrast. HBP increased steadily until it peaked on day 5 and then dropped on day 8.

**Figure 3.**
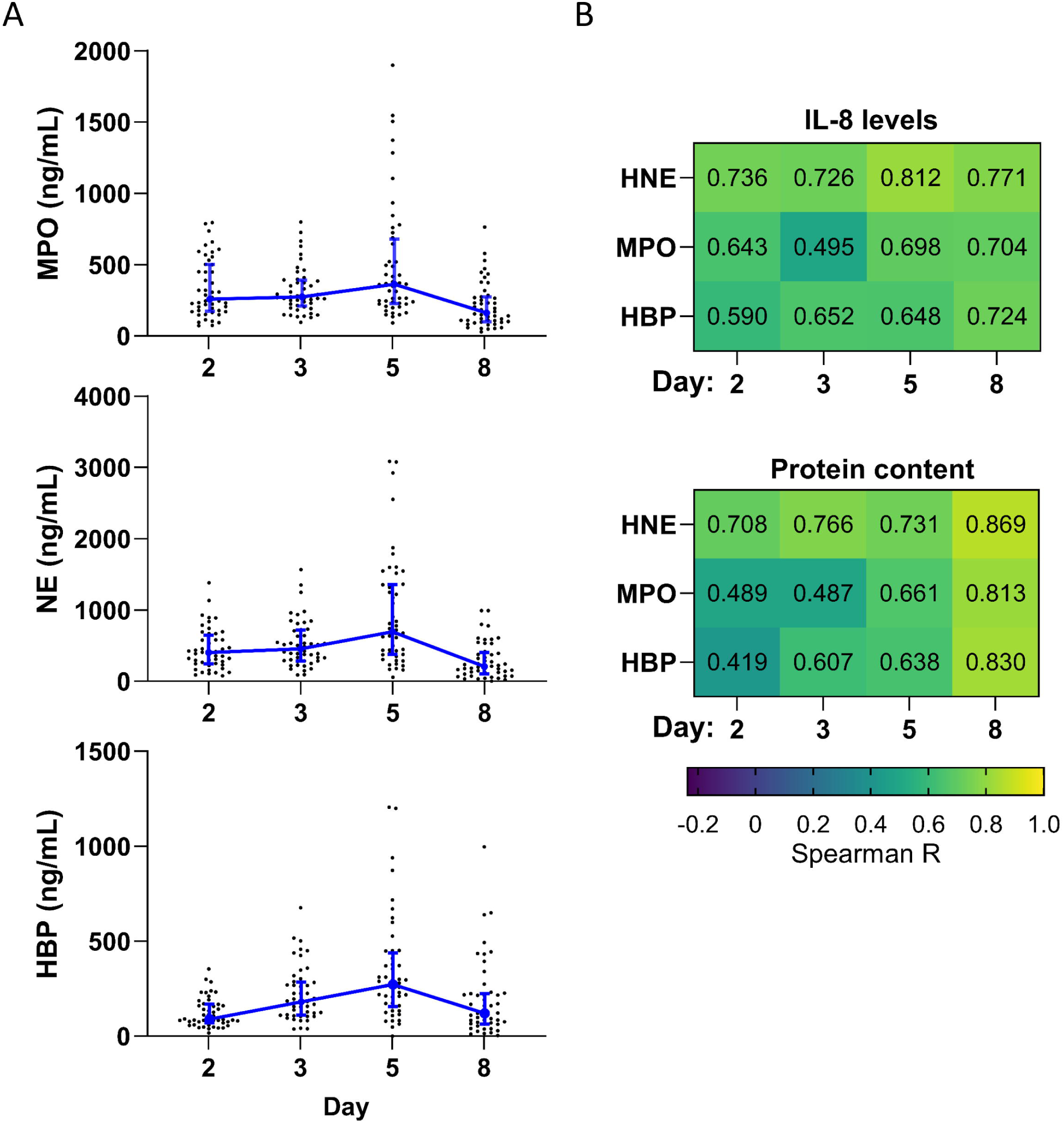
Neutrophil proteins over time. A) Levels of three different neutrophil proteins measured in dressing fluid over time. Each data point represents the concentration of a specific neutrophil protein in a single wound, while the blue line is the median of all wounds that day (n = 48), and whiskers are the interquartile range. B) Spearman correlation coefficients comparing levels of each neutrophil protein with IL-8 (top) and protein content (bottom) (n = 48).

The levels of each neutrophil protein were moderately to strongly correlated with levels of IL-8, a potent chemoattractant for neutrophils, on each day (R = 0.495 to 0.812) (Figure 3B). Neutrophil recruitment and activation are associated with neutrophil extravasation from the blood to the tissues and consequent capillary leakage. We thus determined the correlation coefficient between the level of each neutrophil protein and protein content of the dressing fluid (a proxy for exudation) (Figure 3B) and found that there was a moderate to strong correlation between neutrophil protein levels and protein content on all days (R = 0.419 to 0.869), particularly on day 8 (R = 0.813 to 0.867). Notably, this correlation was stronger than it was for most cytokines and occurred later (Figure 2D vs Figure 3B), suggesting that inflammation and neutrophil recruitment/activation may drive exudation in different stages of wound healing.

To confirm that measured neutrophil protein levels are not dependent on the amount of protein content in the dressing fluid, we normalized the levels of each neutrophil protein for the protein content of that same sample (Supplementary Figure 3B). The overall dynamics changed slightly, with normalized neutrophil proteins reaching their peak level on day 3, instead of day 5.

### Bacterial levels

To explore how levels of aerobic cultivable bacteria in normally healing epidermal wounds change over time, we quantified the number of bacteria in swab and dressing fluid extract samples. Immediately after formation of the wound (Day 1), detected bacterial levels were zero in most of the swab samples (Figure 4A, Supplementary Table 1). Subsequent samples revealed the rapid reconstitution of cultivable bacteria over time already one day after the wounds were formed (Day 2), corresponding to a median of 2.9×10^3^ CFU per swab and 1.9×10^4^ CFU per dressing. Thereafter bacterial levels in the wound increased steadily until day 8 when they reached a peak, corresponding to a median of 1.7×10^6^ CFU per swab and 1.6×10^7^ CFU per dressing. On day 11, bacterial levels decreased slightly in swabs to a median of 1.3×10^6^ CFU per swab. They decreased to a much larger extent in dressings on day 11, to a median of 1.4× 10^6^ CFU per dressing.

**Figure 4.**
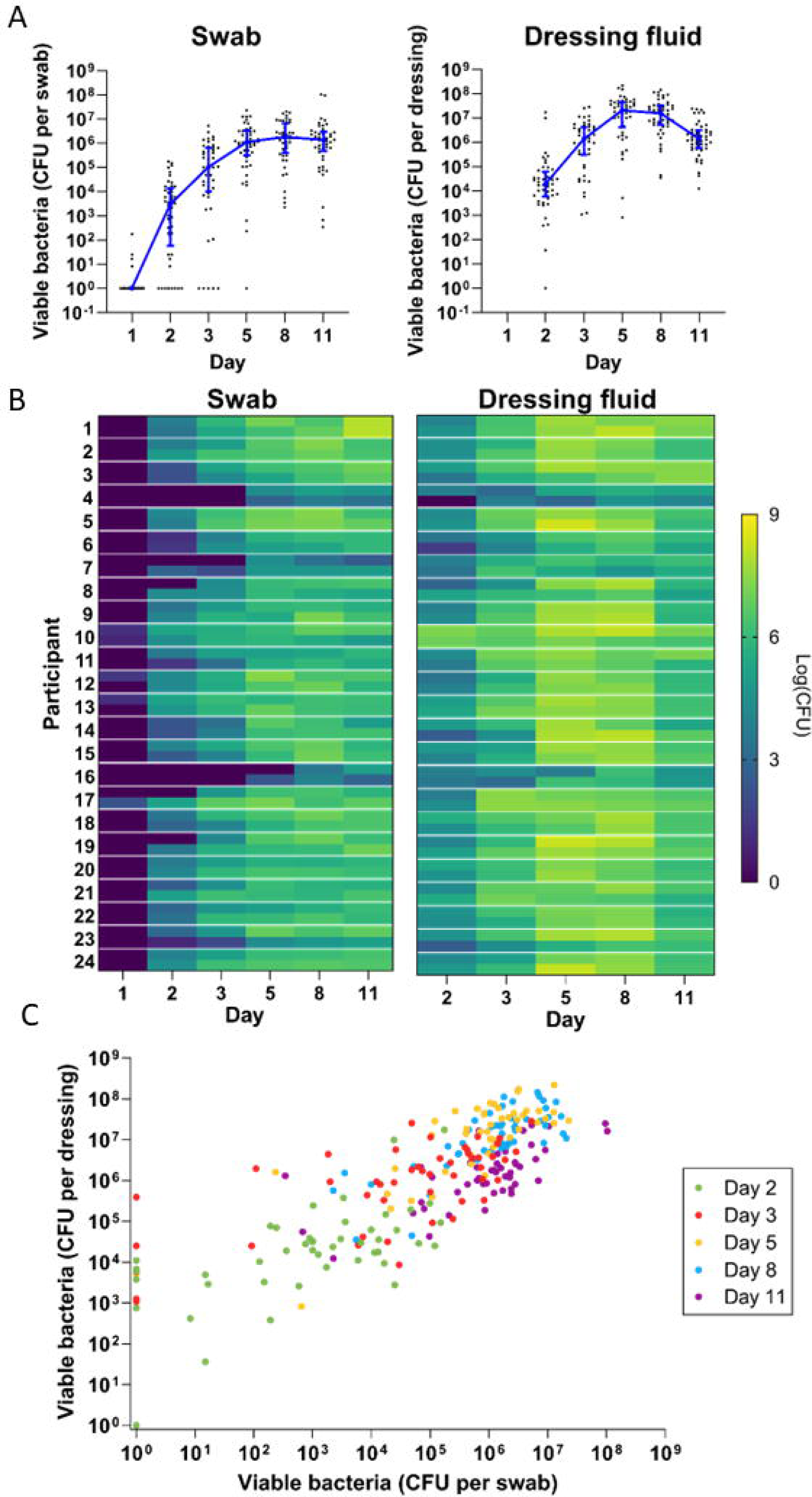
Bacterial development over time. A) Quantitative bacterial counts in swab samples (left) and dressing fluid samples (right) over time. Each data point is the bacterial level measured in a single wound, while the blue line is the median of all wounds that day (n = 48) and whiskers are the interquartile range. Zero values were replaced with 1 in the graph to enable visualization on the logarithmic axis. B) Heat map of quantitative bacterial counts in swab samples (left) and dressing fluid samples (right) from each wound. Data from the left and right wound are each presented in separate cells at the top and bottom of each row, respectively. C) Scatterplot showing quantitative bacterial counts in swabs vs quantitative bacterial counts in dressing fluid in each wound. Zero values were replaced with 1 in the graph to enable visualization on the logarithmic axis. Each day is indicated in a different colour.

Heat maps of bacterial levels in each wound showed considerable individual variation (Figure 4B). Generally, wounds with low bacterial levels in swabs also appeared to have low bacterial levels in dressing fluids, and vice versa. To confirm this, we plotted bacterial levels in swabs and dressings as a scatterplot (Figure 4C) and determined the Spearman correlation coefficient on each day and overall (Table 2). The data indicated a strong correlation between bacterial levels in swab and dressing fluid samples on each day (R = 0.607 – 0.705) and overall (R = 0.794).

**Table 2.** Correlation coefficients for bacterial counts in various samples per day and overall.

| Comparison | Day | Spearman R | P value | 95% confidence interval |
| --- | --- | --- | --- | --- |
| CFU in swabs (left leg vs right leg) | Day 2 | 0.368 | 0.077 | -0.0545 to 0.678 |
|  | Day 3 | 0.380 | 0.067 | -0.0402 to 0.686 |
|  | Day 5 | 0.489 | 0.015 | 0.0937 to 0.751 |
|  | Day 8 | 0.511 | 0.011 | 0.124 to 0.764 |
|  | Day 11 | 0.305 | 0.147 | -0.124 to 0.639 |
|  | Overall | 0.723 | <0.0001 | 0.621 to 0.801 |
| CFU in dressing fluid (left leg vs right leg) | Day 2 | 0.230 | 0.279 | -0.202 to 0.588 |
|  | Day 3 | 0.653 | 0.001 | 0.328 to 0.840 |
|  | Day 5 | 0.444 | 0.030 | 0.037 to 0.725 |
|  | Day 8 | 0.237 | 0.266 | -0.200 to 0.592 |
|  | Day 11 | 0.597 | 0.002 | 0.243 to 0.810 |
|  | Overall | 0.816 | <0.0001 | 0.744 to 0.870 |
| CFU in swabs vs CFU in dressing fluid | Day 2 | 0.661 | <0.0001 | 0.457 to 0.800 |
|  | Day 3 | 0.617 | <0.0001 | 0.397 to 0.770 |
|  | Day 5 | 0.660 | <0.0001 | 0.456 to 0.800 |
|  | Day 8 | 0.705 | <0.0001 | 0.520 to 0.827 |
|  | Day 11 | 0.607 | <0.0001 | 0.383 to 0.764 |
|  | Overall | 0.794 | <0.0001 | 0.741 to 0.838 |
| Protein content in dressing fluid vs CFU in swabs | Day 2 | 0.0490 | 0.741 | -0.247 to 0.336 |
|  | Day 3 | 0.120 | 0.414 | -0.178 to 0.398 |
|  | Day 5 | 0.165 | 0.262 | -0.133 to 0.436 |
|  | Day 8 | 0.221 | 0.132 | -0.0763 to 0.482 |
|  | Day 11 | 0.0550 | 0.710 | -0.241 to 0.342 |
|  | Overall | -0.119 | 0.0658 | -0.246 to 0.0115 |

CFU = colony forming units

Bacterial levels plotted on the heatmap appeared to be similar in left and right wounds in most participants, but with some variation. To confirm this, we determined the Spearman correlation coefficient on each day and overall (Table 2). The data indicated a weak to moderate correlation between left and right wounds on each day in swabs (R = 0.305 – 0.511) and dressings (R = 0.230 – 0.653), but a strong correlation overall in both swabs (R = 0.723) and dressings (R = 0.816) when all days were included, indicating that there is moderate variation between left and right wounds in each individual as the wound heals, but that overall bacterial levels do not differ greatly between wounds on the left and right leg, corroborating the data presented in Figure 4C and Table 2, which showed no difference in median bacterial levels between wounds on the left and right leg.

### Identity of bacteria

To examine the major cultivable bacterial species that colonize wounds during the healing process we analyzed bacterial colonies that grew from streaks of each swab and dressing sample on day 3 and 8. Overall the identified bacterial species were dominated by commensal species, such as *S. epidermidis*, which was found in swab and dressing samples from the majority of wounds on day 3 and 8 and in swabs and dressings (Table 3, see also Supplementary Figure 2 for a complete representation of the identified bacteria). Some wounds also contained *S. aureus* and *Bacillus sp.*, which can sometimes be pathogenic, though no signs of infection were observed in these wounds (Supplementary Figure 4). On day 8, we observed an increase in the number of colonies identified as the potential pathogen *S. aureus* and the commensal *Corynebacterium sp* (Supplementary Figure 2). On examination of images of the wounds over time we noted that, in spite of differences in the cultivable bacteria at the wound surface and in dressings, all wounds healed at roughly the same pace and had a similar appearance at each stage ^29^.

**Table 3.**
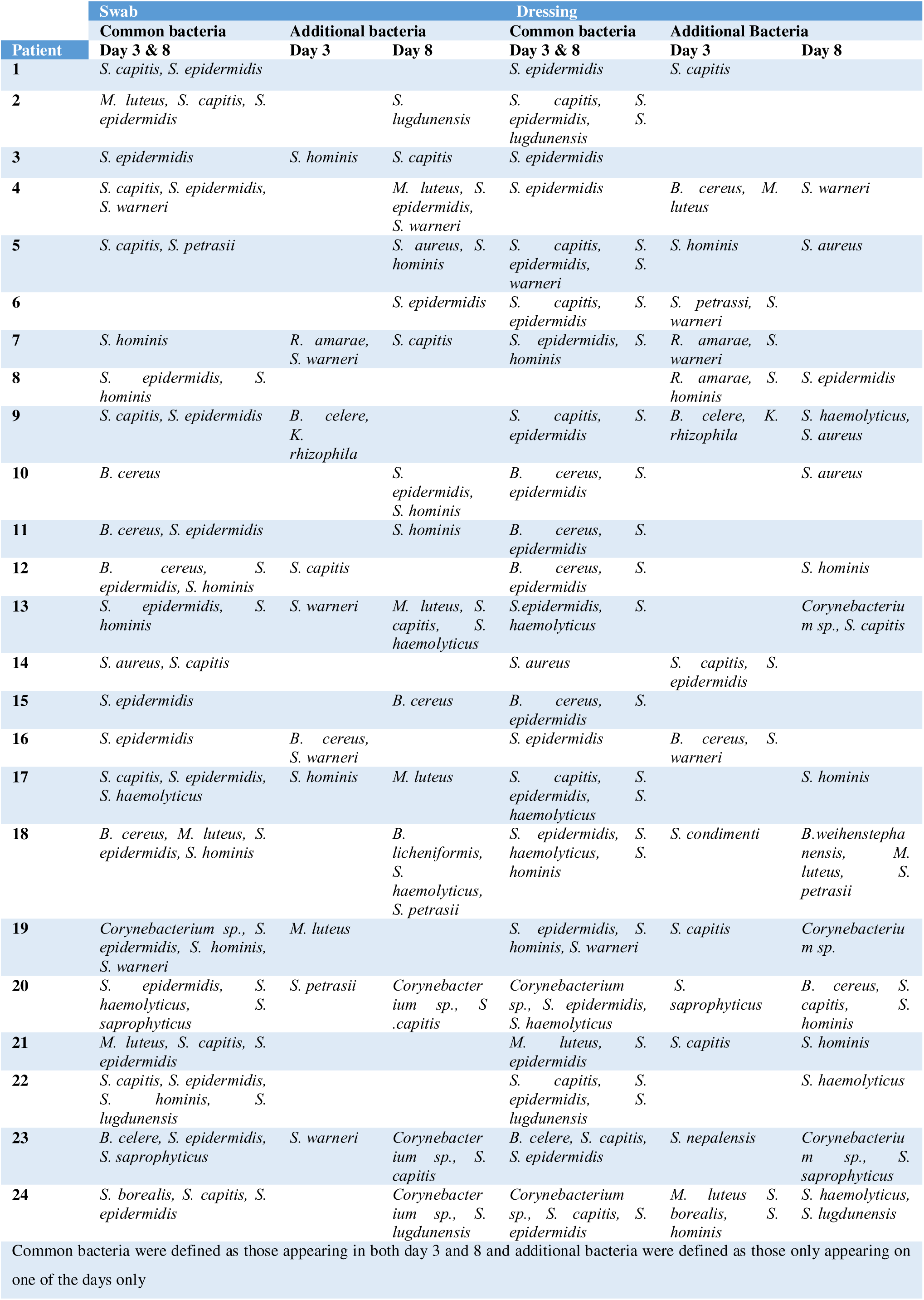
Comparison of similar MALDI TOF identified bacterial species during study day 3 and day 8 for both swab and dressing fluid samples.

### Correlation between inflammation, exudation, neutrophil proteins, and bacterial levels

To determine whether the observed bacterial levels affect inflammation, we determined the correlation coefficient between the level of each cytokine and the bacterial level detected in swabs in that wound and plotted the resulting correlation coefficients as a heatmap (Figure 5A). For most cytokines, correlations with bacterial counts were minimal or absent on days 2, 3, and 8, with only a weak correlation observed on day 5. However, certain cytokines showed moderate correlations with bacterial counts, including TNF-α on day 3 and 5 (R = 0.475 and 0.488 respectively), IL-10 on day 5 (R = 0.531), and IL-1β on days 5 and 8 (R = 0.497 and 0.480 respectively). The correlation coefficients increased slightly for most cytokines when they were normalized for protein content (Supplementary Figure 3C). In contrast, we found little to no correlation between bacterial levels in swabs and neutrophil proteins on each day (R = 0.011 to 0.353).

**Figure 5.**
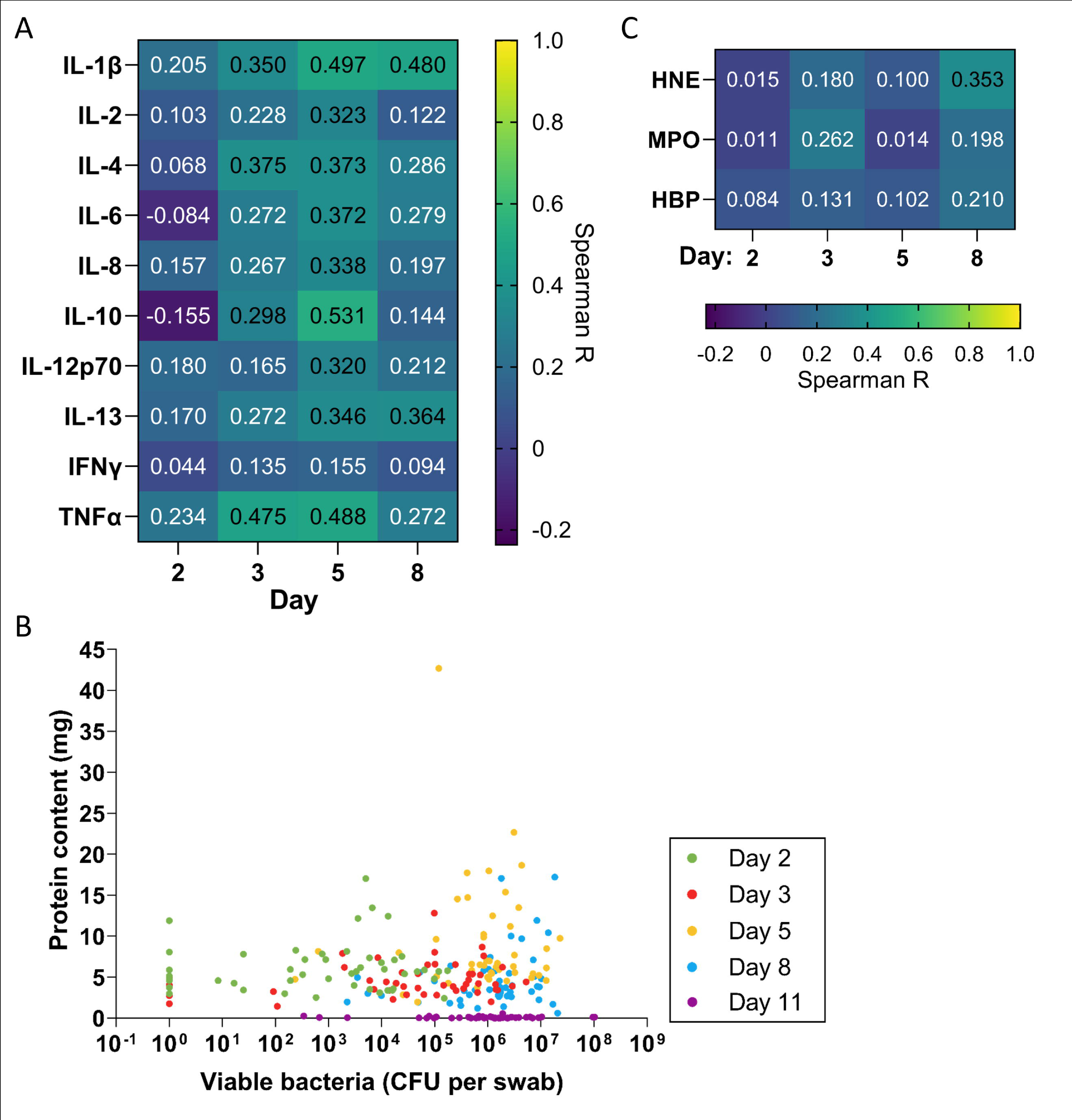
Correlation of inflammation, exudation and neutrophil proteins with bacterial levels. A) Spearman correlation coefficients comparing levels of each cytokine with quantitative bacterial counts measured in swabs (n = 48). B) Scatterplot comparing protein concentration in dressing fluids vs quantitative bacterial counts measured in swabs from each wound. Zero values were replaced with 1 in the graph to enable visualization on the logarithmic axis. C) Spearman correlation coefficients comparing levels of each neutrophil protein with quantitative bacterial counts measured in swabs (n = 48).

To explore whether the observed level of viable bacteria detected on the wound surface affects the level of exudation, we plotted bacterial levels measured in swabs against protein content as a scatterplot (Figure 5B) and determined the Spearman correlation coefficient on each day and overall (Table 2). The data indicated that there is weak to no correlation between bacterial levels and protein content on each day (R = 0.049 – 0.221) and no correlation overall (R = –0.119). In addition to the bacteria directly on the wound surface, bacteria in the dressings could contribute to the induction of inflammation, potentially releasing various proinflammatory products such as lipoteichoic acid (LTA) and peptidoglycan. Thus, we examined the correlation of cytokine levels and neutrophil proteins with bacterial counts in dressing fluid (Supplementary Figure 5A). In general, the correlations between cytokines and dressing bacterial loads were similar to their correlation with bacterial levels measured in swabs, with slightly higher correlations observed for TNF-α, IL-10 and IL-1β at the peak of inflammation on day 5 (Supplementary Figure 5 A and B).

## Discussion

In this study, we comprehensively describe the dynamics of exudation, inflammation, neutrophil proteins, and bacteria in epidermal suction blister wounds and their interrelatedness during the wound healing process (Figure 6). Inflammation plays a key role in the wound healing process. To assess inflammation over time in normally healing epidermal wounds, we measured levels of 10 cytokines in dressing fluid on days 2, 3, 5, and 8. While cytokine levels varied, they showed similar overall trends: most cytokines stayed constant from days 2 to 3, though some (IL-4, IL-6, IL-10) declined slightly. IFN-γ, however, increased steadily until day 5. All cytokines peaked on day 5 before decreasing on day 8.

**Figure 6.**
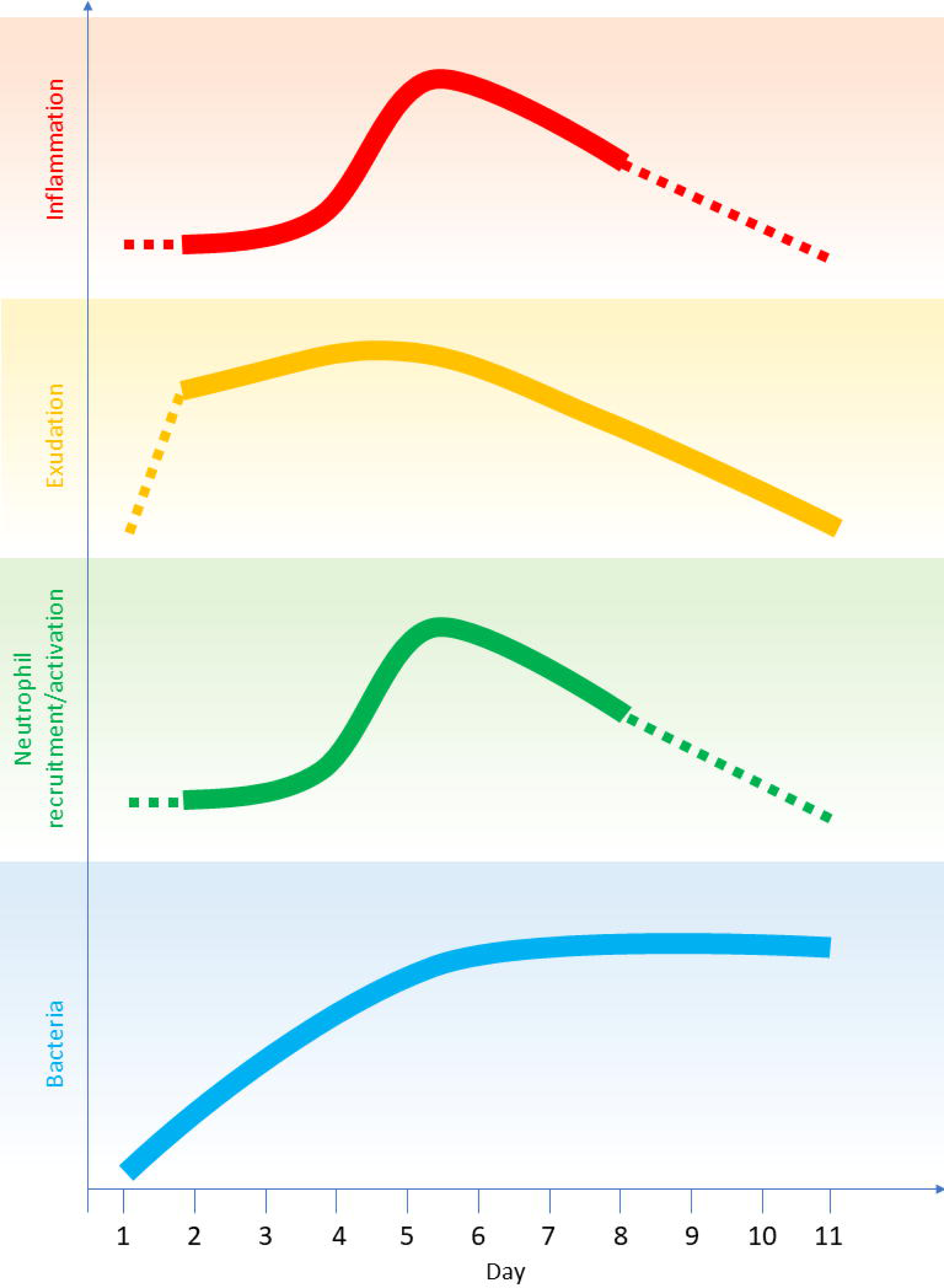
Overview of the dynamics of bacterial levels, exudation, neutrophil proteins, and inflammation in normally healing wounds over time, and the relationship between them. Dashed lines are a reasonable extrapolation to timepoints not measured in this study.

Inflammation also plays a role in driving wound exudation, with inflammatory cytokines contributing to this process. Therefore, as exudate contains plasma-derived proteins like albumin, expressing cytokine levels in dressing fluid relative to total protein can introduce a confounding factor. As exudation increases, the dilution of cytokines by the influx of plasma proteins can obscure the actual amounts of cytokines released in the wound. To mitigate this effect, we present cytokine levels in two complementary ways: per mL to reflect their absolute concentration, and per mg of total protein to account for variations in protein content due to exudation. Using both approaches provides a nuanced representation of cytokine dynamics and strengthens the conclusions of the analysis. We found that the overall dynamics of the cytokines remained consistent even after normalization for protein content, indicating that the trends observed were not significantly influenced by exudate dilution. This suggests that the inflammatory response, as reflected by cytokine levels, is a robust process that correlates with wound healing progression independently of variations in exudate volume. Overall, our findings are in line with several studies that have explored cytokine dynamics in wound healing, emphasizing the inflammatory phase and its downregulation^5,7,8^, highlighting the importance of a balanced inflammatory response for wound healing.

Wound exudation is a critical aspect of the wound healing process ^6^. We found that exudation, as indicated by the level of protein content in dressing fluid, was moderately high already on day 2, then peaked on day 5, before declining on day 8. Inflammation, as indicated by cytokine levels, was correlated with protein content in dressing fluid, particularly on day 5 when inflammation was highest, corroborating the link between inflammation and capillary leakage, which in turn leads to exudation ^6^.

Neutrophils play a crucial role in wound healing and bacterial control and both neutrophil infiltration and activation are known to increase in the inflammatory stage of wound healing ^3–5^. We observed that neutrophil protein levels, which could be attributed to increased neutrophil recruitment and activation, followed similar dynamics to inflammation. MPO and NE increased on day 5, while HBP began to increase already on day 3. These dynamics are consistent with the fact that HBP is stored in the secretory vesicles which have a high propensity for release, while MPO and NE are stored exclusively in the azurophilic granules which have a low propensity for release ^33^. Neutrophil protein levels were strongly correlated with IL-8, a potent chemoattractant for neutrophils.

A novel observation in this study is identification of HBP in epidermal wounds.^34^ HBP is known to increase vascular permeability, leading to capillary leakage and subsequent exudation. This mechanism has been observed in various conditions, including burn wounds and sepsis. For instance, studies have shown that HBP, released from activated neutrophils, is increased in severe burn injuries^35^ as well as in non-healing venous wounds^36^. Similarly, in sepsis, elevated levels of HBP are associated with increased endothelial permeability, leading to fluid leakage and organ dysfunction^37^. Therefore, HBP’s ability to increase vascular permeability may explain the correlation between HBP and wound fluid protein levels (exudation) observed in our study, highlighting a novel link between HBP and exudation in epidermal wound healing.

The understanding that bacteria, particularly various staphylococcal species, can play a role in modulating normal wound healing^15–17,38–41^ guided our focus on viable aerobic bacteria in this study. While this approach naturally limits the detection scope compared with broader methods like amplicon or metagenomic sequencing—capable of identifying anaerobes and non-culturable organisms—it provides a targeted and clinically relevant analysis of the growth of these key commensals. This is particularly important as, unlike bacterial DNA, the presence of viable bacteria reflects bacterial metabolic activity, which could have direct implications for wound healing dynamics. In addition, the skin microbiome is unique in its composition and activity compared with other human microbiomes. Notably, skin has a lower proportion of viable bacteria, meaning that a significant fraction of bacterial DNA present on the skin surface may come from non-viable organisms. Traditional sequencing methods, such as 16S rRNA gene sequencing, can overestimate the richness and diversity of the skin microbiome by detecting DNA from dead or inactive bacteria^42^, which do not influence biological processes of wound healing, for example by producing various proteases^43^. This distinction between live and dead bacterial populations is crucial when studying the impact of commensals like *S. epidermidis* on healing. Indeed, the repopulation of the skin microbiome after a disturbance is driven by an underlying viable bacterial population^42^. In this context, the focus on viable, cultivable bacteria allowed us to gain insight into the growth kinetics of commensal species like *S. epidermidis* during normal wound healing.

At the onset of wound formation, minimal amounts of cultivable bacteria were observed on the wound surface, likely due to the removal of the epidermis and prior skin disinfection with ethanol. Shortly after wounding, bacterial colonization was detected, with levels progressively increasing until day 8, followed by a slight decline by day 11. Although there was some variation in the patterns of cultivable bacteria between individuals, *S. epidermidis* was consistently dominant, and most wounds contained bacteria typically classified as skin commensals. This finding aligns with the view that commensals are conducive to normal healing processes^38–41,44,45^.

Notably, consistent with recent findings on the pro-inflammatory roles of commensal bacteria^16^, we observed a moderate correlation between bacterial levels on day 5 and the pro-inflammatory cytokines TNF-α and IL-1β, suggesting that bacteria can influence inflammation even in normally healing epidermal wounds. Interestingly, a similar correlation was found with IL-10, an anti-inflammatory cytokine known to promote a regenerative phenotype in experimental dermal wounds^46^. It is also important to note that several other cytokines showed little to no correlation with bacterial levels, underscoring the complex, and likely selective, cytokine responses induced by bacteria in human epidermal wounds

Interestingly, previous studies on wound complications in skin grafting procedures in patients have indeed shown that elevated bacterial levels, largely consisting of commensals, are associated with wound complications^47,48^, including increased inflammation reflected by elevated levels of TNF-α and IL-1β^49^. These findings underscore the dual role of commensals in both facilitating and, under certain conditions, potentially hindering wound healing. It should also be noted however that some wounds harbored bacteria that sometimes can act as pathogens, such as *S. aureus*, which often underlies postoperative infections ^50^. However it is also worth noting that the presence of *S. aureus* does not define infection *per se*, as the bacterium can, like other commensals, also occur in normally healing wounds ^49^. Moreover, it has been shown that, like commensals, *S. aureus* can enhance skin regeneration through IL1β-keratinocyte-dependent IL1R/MyD88 signaling ^12^.

Taken together, these findings collectively highlight the highly dynamic nature of the wound healing process. Bacterial colonization, inflammation, neutrophil activity, and exudation appear to be interrelated but not solely dependent on one another (Figure 6). From a methodological perspective, our study also revealed a strong correlation between observed levels of the studied bacteria in swab and dressing fluid samples on each day and overall. Given this correlation, analysis of dressing fluid bacteria could be an alternative way to collect information about bacteria in the wound environment. Furthermore, while intra-individual variation existed between left and right wounds in each participant, shown by the poor correlation between bacterial levels in wounds on the left and right leg, overall bacterial dynamics followed the same trend on both legs.

A key strength of this study is the use of a suction blister model, which allowed for the creation of standardized wounds in human subjects and facilitated a detailed, multifaceted analysis of bacterial dynamics and host factors throughout the healing process. However, there are limitations to consider. First, the bacterial levels observed here may not be representative of bacterial levels in other wound types, particularly since the occlusive dressings and hydrogel used in this study could have enhanced the moisture in the wound environment, potentially influencing bacterial growth ^51–53^. Moisture is a well-known factor for bacterial proliferation, as occluded skin displays a significant increase in bacterial numbers after just one day ^52^. It is therefore possible that the moist environment under the occlusive polyurethane-based dressing used here promoted bacterial growth compared to intact skin, which naturally harbors fewer bacteria ^54,55^. Our choice of dressings may have thus affected the observed bacterial dynamics. As re-epithelialization progressed and exudation diminished, the decrease in bacterial levels by day 11 could possibly reflect both reduced access to nutrients in wound fluid and a shift in the host immune response. Moreover, the wounds in this study received a hydroxyethyl cellulose (HEC)-based gel, which added additional moisture. HEC is widely recognized for its high biocompatibility and low immunogenicity^56^, ensuring that it did not introduce any confounding effects beyond moisturizing the wound, and similar inert hydrogels are often used in the clinic. Thus, these results represent a clinically relevant scenario, where moisture contributes to the overall dynamics of bacterial colonization.

Finally, as discussed above, culture-based methods can underestimate bacterial presence, meaning the total bacterial burden (including viable and non-viable bacteria) in these wounds could be higher than reported. Future studies could address these limitations by exploring alternative methods for deeper microbial sampling and/or utilize amplicon or metagenomics-based approaches to study the complete microbiome.

### Conclusions

In conclusion, this study highlights the complex interplay between inflammation, exudation, neutrophil activity, and bacterial colonization in normal epidermal wound healing. Cytokine levels peaked on day 5, mirroring neutrophil-derived proteins such as MPO, NE, and HBP. The correlation between neutrophil chemoattractant IL-8 and neutrophil activity underscores the role of neutrophils in this process. Additionally, the identification of HBP – a key player in promoting vascular permeability and exudation – offers novel insights into its role in wound healing. The dynamics of bacteria, such as *S. epidermidis*, further underline the presence of commensals in normal wound healing, while possible relationships between bacterial levels and the cytokines TNF-α, IL-1β, and IL-10 point to the potential for bacterial influence on inflammation in some contexts.

## Data Availability

The data that support the findings of this study are openly available online in Zenodo at http://doi.org/10.5281/zenodo.10283373, reference number 10283373.

## Acknowledgments

Xinnate AB provided the funding, project management resources, and expertise for the regulatory development, enabling the safety study that generated the control samples used in this work. We would like to thank Dr. Andreas Sonesson for valuable discussions and Susanne Erdmann and Anne Nielsen and other personnel at the Department of Dermatology Lund. Lastly, we would also like to thank Åsa Håkansson and Kerstin Weber and other personnel at the Clinical Trial Unit at Skåne University Hospital Lund for support and use of their facilities during the clinical study.

## Sources of funding

The exploratory study presented here was supported by grants from the Swedish Research Council (project 2017-02341, 2020-02016), Edvard Welanders Stiftelse and Finsenstiftelsen (Hudfonden), the Royal Physiographic Society, the Crafoord and Österlund Foundations, and the Swedish Government Funds for Clinical Research (ALF).

## Authors’ contributions

The following authors contributed to each of the following roles (defined according to the CRediT taxonomy): Conceptualization, SL, GP, KW, MP, KS, AS; design of methodology, SL, KW, MP; formal analysis (statistics), GP, JFPC, CL, JF; investigation (performing experiments and data collection) and validation, SL, GP, KW, JFPC, ACS, FF, CL, BN; resources (provision of patients and samples), SL, KW, KS, AS; data curation, JFPC, JF; writing (original draft preparation), SL, KW, JF, AS; writing (review and editing), SL, GP, KW, JFPC, ACS, FF, CL, BN, EH, JF, MP, KS, AS.; visualization, GP, JFPC, CL, EH, JF; supervision, GP, KS, AS; project administration, SL, GP, KW, JF, AS; funding acquisition, AS. All authors have read and agreed to the published version of the manuscript.

## Data sharing and availability

The data that support the findings of this study are openly available in Zenodo at http://doi.org/10.5281/zenodo.10283373, reference number 10283373.

## Conflict of Interest disclosure statement

AS is a founder of in2cure AB, a parent company of Xinnate AB which was the sponsor of the clinical trial from which the biobank samples used in this study are derived. GP is employed part-time (20%) by Xinnate AB. JF provides consulting services to Xinnate AB. The other authors have declared that no conflict of interest exists.

## Transparency statement

The lead author (manuscript guarantor) affirms that this manuscript is an honest, accurate, and transparent account of the study being reported; that no important aspects of the study have been omitted; and that any discrepancies from the study as planned (and, if relevant, registered) have been explained.

This study was previously made available as a preprint at https://doi.org/10.1101/2023.12.07.23299659

## Abbreviations

BCA: bicinchoninic acid
CFU: colony forming units
ELISA: enzyme-linked immunosorbent assay
HBP: heparin-binding protein
IFN: interferon
IL: interleukin
IL-1R: Interleukin-1 receptor
MALDI-TOF: Matrix Assisted Laser Desorption lonization -Time Of Flight
MBT: MALDI Biotyper
MPO: myeloperoxidase
MS: mass spectrometry
MyD88: Myeloid differentiation primary response 88
NE: neutrophil elastase
PBS: phosphate-buffered saline
TCP: thrombin-derived C-terminal peptide
TNF: tumor necrosis factor

## Notes

### Competing Interest Statement

A.S. is a founder of in2cure AB, a parent company of Xinnate AB which was the sponsor of the clinical trial from which the biobank samples used in this study are derived. G.P. is employed part-time (20%) by Xinnate AB. The other authors have declared that no conflict of interest exists.

### Clinical Trial

NCT05378997

### Funding Statement

This study was funded by grants from the Swedish Research Council (project 2017-02341, 2020-02016), Edvard Welanders Stiftelse and Finsenstiftelsen (Hudfonden), the Royal Physiographic Society, the Crafoord and Österlund Foundations, and the Swedish Government Funds for Clinical Research (ALF).

### Author Declarations

The Swedish ethical review authority (etikprövningsmyndigheten) gave ethical approval for this work (application number 2022-00527-01).

### Summary of Updates

We have clarified that our primary objective was to investigate inflammatory biomarkers and selected neutrophil proteins, along with utilizing a targeted MALDI-TOF-based methodology for bacterial identification. We corrected our terminology to "cultivable bacteria" instead of "microbiome" and emphasized our focus on inflammatory markers including cytokines and neutrophil proteins, exudation and protein content, as well as bacterial dynamics. We justified using Todd-Hewitt (TH) agar for bacterial quantification, explaining its suitability and limitations, which are now included in the discussion. Our colony selection method for MALDI-TOF analysis has been clarified, ensuring a representative sample based on morphology. We clarified the inclusion criteria for subjects and control wounds, specifying the use of hydroxyethyl cellulose (HEC) gel and discussing its potential effects. We detailed the sample processing time and provided data showing bacterial viability was not affected, as explained in the methods section. We included a reference for the swabbing procedure and explained why direct application of swabs or dressings to culture media was not feasible. Our choice of blood agar was explained along with its limitations, and the methodology for colony selection was outlined to address concerns about missing microorganisms with similar morphology. Finally, we clarified that the presence of S. aureus did not indicate infection, as it can be part of the commensal skin flora, and included this in the discussion.

