## supplement for "Temporal Dynamics and Interrelations of Cytokines, Neutrophil Proteins, Exudation, and Bacterial Colonization in Epidermal Wound Healing"

### Supplementary methods

#### Swab procedure

To collect swab samples from wounds, the swab was pre-wetted with sterile phosphate-buffered saline (PBS). Using a twisting motion, the swab was rotated 10 times using light pressure, covering the entire wound and 2 mm outside of the initial wound edge on each side, and then placed into a microfuge tube with 0.5 mL sterile PBS and kept on ice. The tubes were vortexed to dislodge bacteria from the swab. In total, swabs and extracted samples were on ice for no more than 2 hours prior to quantification. We have verified that keeping the samples on ice for up to 24 hours on ice does not affect viability of the bacteria analyzed (Supplementary Figure 1B).

#### Dressing extraction

placed into an empty 5 mL tube, and kept on ice. Each dressing was weighed in the tube, and then the dressing was removed and placed into a 5 mL syringe fixed with a stopper. Two milliliters of cold, sterile 10 mM Tris buffer at pH 7.4 was added to the syringe, and the syringe was vortexed for 5 minutes. The fluid was extracted from the dressing by depressing the plunger, emptying the contents into the original 5 mL collection tube in which the dressing had originally been placed. The extracted fluid was kept on ice. We added 100× Halt Protease Inhibitor Cocktail (Thermo Fisher Scientific, USA) to half of the dressing fluid per the manufacturer’s directions to a final strength of 1×. Samples were then aliquoted and frozen at –80°C. In total, dressing and extracted samples were on ice for no more than 2 hours prior to quantification and storage.

*Calculation of dressing fluid extract volume*

To calculate the average weight of the dressing and the tube, six unused dressings and six unused 5 mL collection tubes were weighed, and together were found to have an average weight of 3285.4 mg. This weight was subtracted from the recorded weight of each dressing sample (which had been weighed in the collection tube as described above). We assumed that wound fluid has a density of 1 g/mL and used this to calculate the volume of wound fluid in each dressing sample in milliliters. To calculate the total volume of dressing fluid extract, we added 2 mL to this calculated volume to account for the volume of Tris buffer used during the extraction. This calculated total volume of dressing fluid extract was used to calculate total bacteria and total protein levels as described below.

#### Quantification of cytokines

Cytokines (interferon gamma [IFN]-γ, IL-1β, IL-2, IL-4, IL-6, IL-8, IL-10, IL-12p70, IL-13, tumor necrosis factor [TNF]-α) were measured using a V-PLEX Proinflammatory Panel 1 (human) kit (Meso Scale Diagnostics, Rockville, MD, USA) per the manufacturer’s instructions at a 5× dilution. In some samples IL-8 was above the upper limit of detection and so these were analyzed again using the U-PLEX Biomarker Group 1 (human) kit (Meso Scale Diagnostics) at a 100× dilution. The concentration of each cytokine was calculated using the MSD discovery Workbench analysis software. Samples with a concentration below the detection limit were not able to be quantified by the analysis software and were assigned a value of zero for all analyses. In some analyses, to account for different levels of wound fluid in the dressing extract samples, cytokine levels were normalized for the protein concentration measured by bicinchoninic acid (BCA) assay, as described above, to obtain the amount of each cytokine per mg of protein in the dressing fluid extract sample.

#### Quantification of protein content (wound exudation level)

Dressing fluid was analyzed for protein content by BCA assay (Thermo Fisher, Waltham, MA, USA) per the manufacturer’s directions using bovine serum albumin as the standard. The concentration of protein in mg/mL was calculated from the standard curve. To determine the total protein content in the dressing, this concentration was multiplied by the total volume of dressing fluid extract, calculated as described above.

#### Quantification of neutrophil proteins

Human neutrophil elastase (NE) and human myeloperoxidase (MPO) were measured using ELISA kits from R&D Systems (Minneapolis, MN, USA); heparin-binding protein (HBP) was determined using an ELISA kit from Aviva Systems Biology (San Diego, CA, USA). Samples were analyzed according to the manufacturer’s instructions in duplicate. ELISA signals were read at 450 nm on a microplate reader (BioRad, Hercules, CA, USA). A standard curve was established with reference standards to calculate the concentration in each sample. In some analyses, to account for different levels of wound fluid in the dressing extract samples, neutrophil protein levels were normalized for the protein concentration measured by BCA assay, as described above, to obtain the amount of each neutrophil protein per mg of protein in the dressing fluid extract sample.

#### Quantitative bacterial counts

The swabs and dressing fluid samples were diluted with sterile PBS to generate 7 10-fold serial dilutions (from 10× to 10^7^×). Six separate 10-μL drops of the undiluted sample and each of the dilutions were deposited on a Todd-Hewitt (TH) agar plate. TH is a rich medium that enables identification of a variety of organisms (Supplementary Figure 1A) ^28^ and has the added advantage that it is more translucent than blood agar, which facilitates accurate counts. The plates were incubated at 37°C in 5% CO_2_ overnight. The next morning, the number of colonies was counted and recorded. The number of colony forming units (CFU)/mL was calculated, and then multiplied by the total volume of fluid in the sample (0.5 mL for swabs or the volume calculated from the dressing weight described above for dressing fluid extract) to obtain the total CFU per swab and CFU per dressing.

#### Identification of major cultivable bacteria

Swab and dressing fluid samples were streaked on a blood agar plate and incubated at 37°C overnight To provide a representative overview of the major cultivable bacterial species, six colonies were selected from each plate. If visually distinct colony types were present, at least one of each was chosen based on their relative distribution. If colony morphology was uniform, colonies were selected at random. This approach enabled largely qualitative identification of major bacterial types (Supplementary Figure 2).

Colonies were prepared using the extended direct transfer sample preparation procedure on stainless steel MALDI target plates as described by the manufacturer (Bruker Daltronik GmbH). A Microflex LT/SH SMART MALDI-TOF MS instrument with flexControl v. 3.4 (Bruker Daltronik GmbH) was used to analyze the target plate and collect mass spectra in linear mode over a mass range of 2 to 20 kDa. A spectrum of 240 summed laser shots was acquired for each sample spot. The spectra were analyzed using a MALDI Biotyper (MBT) Compass v. 4.1 with the MBT Compass Library Revision L (DB-9607, 2020) (Bruker Daltronik GmbH). A MALDI Biotyper score of 2.0 or more was required for species level determination, except for *Corynebacterium* where an identification with a score of 2.0 or more was limited to the genus level, and for species belonging to the *Bacillus cereus* complex where the identification was limited to the complex level.

To save the bacterial strains for future use, the sterile toothpick that was used to pick each colony for processing was placed in a sterile tube with freezing medium (Extra broth with 10% glycerol and 20% horse serum) ^33^. This tube was then stored at –80°C until further use. One saved sample of each identified bacterial species was streaked on both a blood agar plate and a TH agar plate and we verified that they exhibit equivalent growth on both types of media (Supplementary Figure 1A).

### Supplementary figures

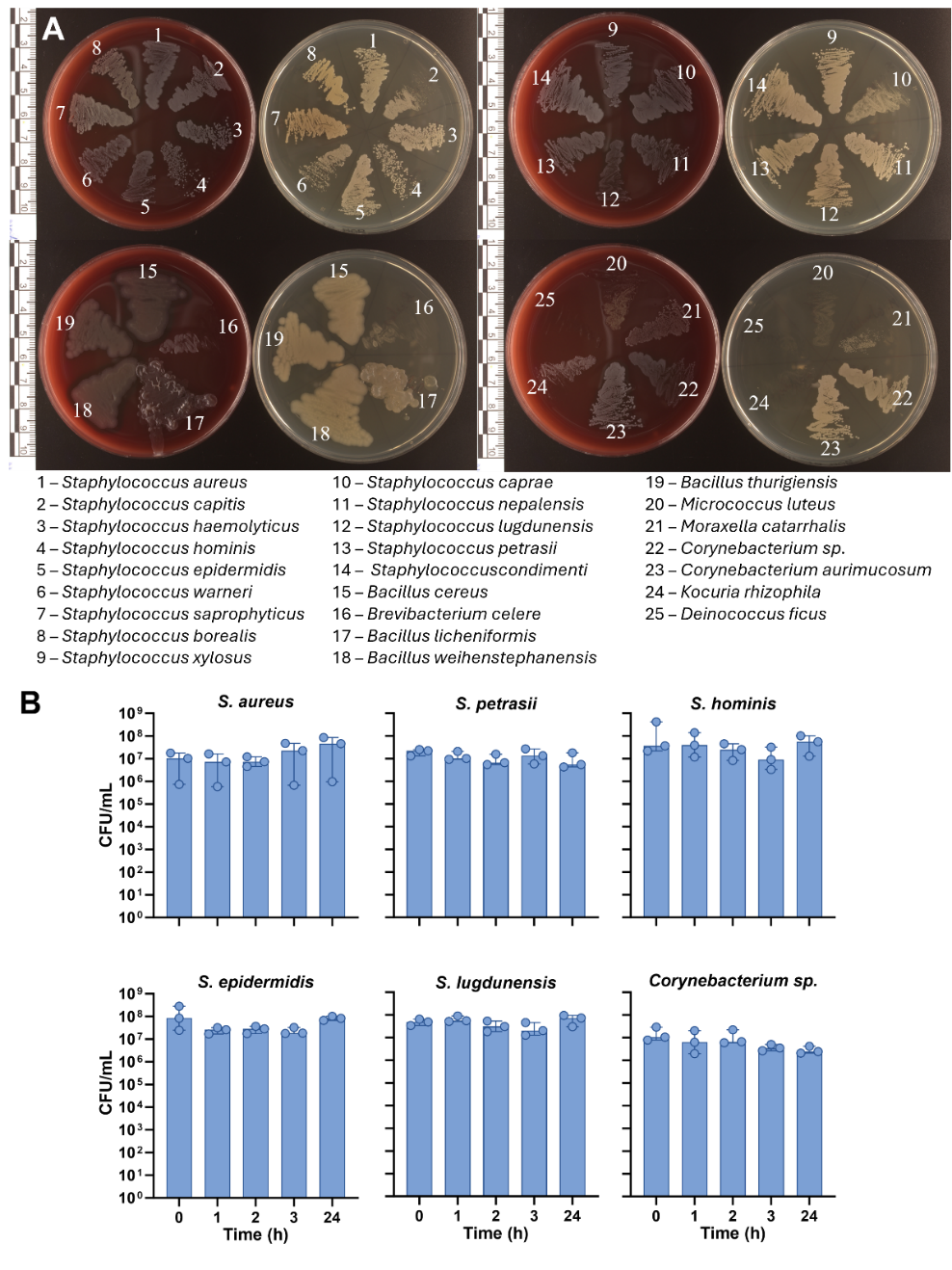
Supplementary Figure 1. **A)** Growth of 25 different bacterial species isolated in the clinical study and identified by MALDI-TOF MS was compared on Todd-Hewitt (TH) and blood agar plates. Bacteria were collected from frozen stocks using a 1 mm inoculation loop and streaked on each plate. Plates were incubated overnight at 37°C in 5% CO_2_. The next morning, plates were imaged using a DSLR camera (Canon EOS Rebel T7i, Japan) equipped with a 60 mm f/2.8 macro lens (Canon EF-S 60 mm, Japan). All tested species except for one – *D. ficus* – grew on both types of media, with some minor differences in terms of colony size, morphology, and relative bacterial amounts.

**B)** The viability of 6 of the bacterial species isolated in the clinical study was determined over time in the same conditions at which the clinical samples were processed. Bacteria were cultured overnight in 5 mL of TH media, centrifuged, and resuspended in PBS to obtain a solution of approximately 1×10^9^ CFU/mL. This solution was absorbed using a cotton swab and then this swab was processed in the same way as the swabs collected from patients, with only the time spent on ice varied between them. Briefly, the swabs were placed in 0.5 mL of sterile PBS and kept on ice for 0, 1, 2, or 3 hours, and then vortexed and plated for quantitative bacterial count as described in the main text. The 0-hour tube was then placed in the refrigerator for 24 hours, and then vortexed and plated for quantitative bacterial count. All samples had similar bacterial loads at each timepoint indicating that bacterial viability is not affected by keeping the samples on ice.

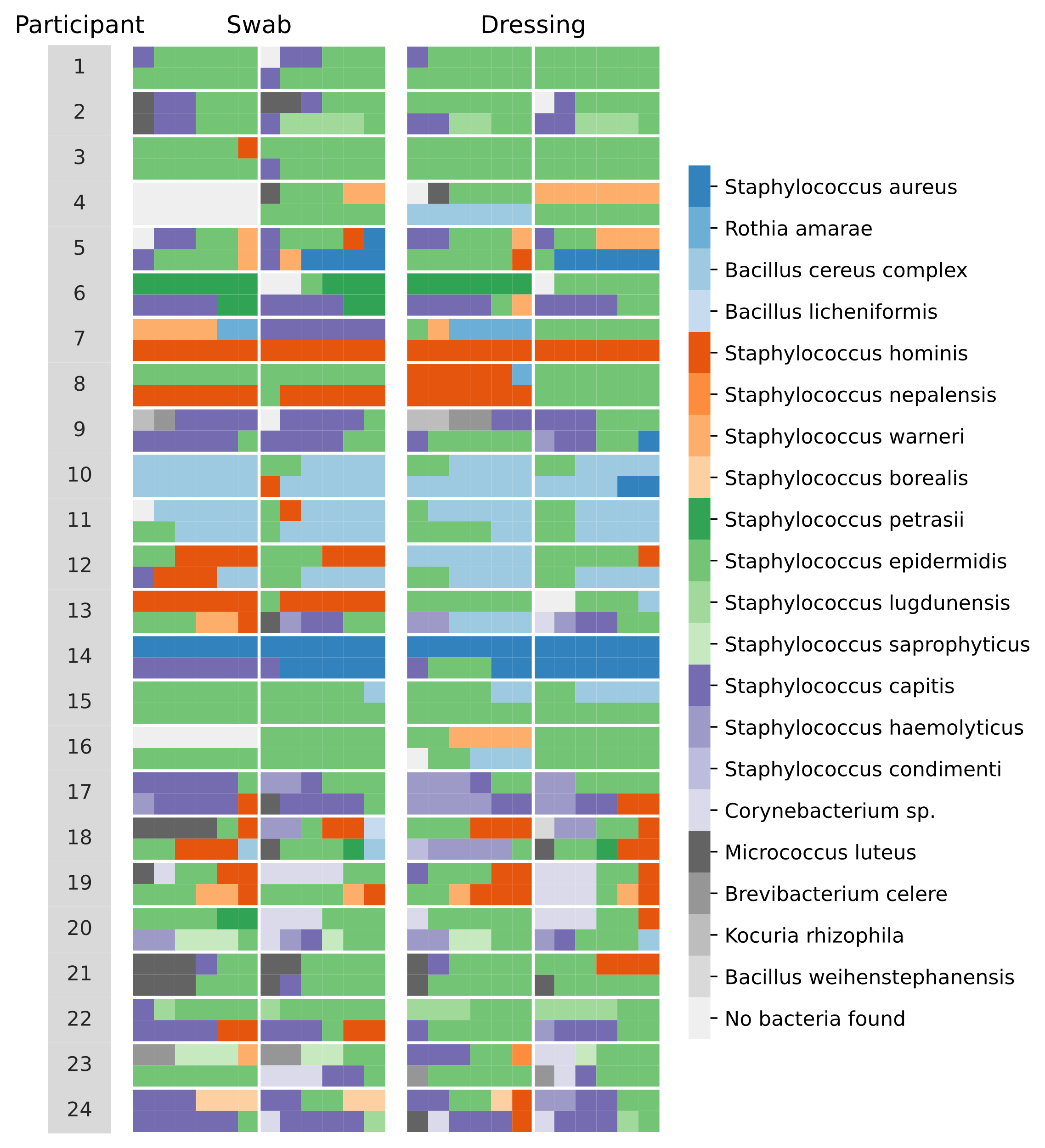

Supplementary Figure 2. **Major cultivable bacterial species identified in wounds on days 3 and 8 post-wounding.** Bacterial species identified by MALDI-TOF MS in six representative colonies picked from swab and dressing fluid cultures from each wound on days 3 and 8. Bacteria from the left wound are in the top cells and bacteria from the right wound are in the bottom cells of each row.

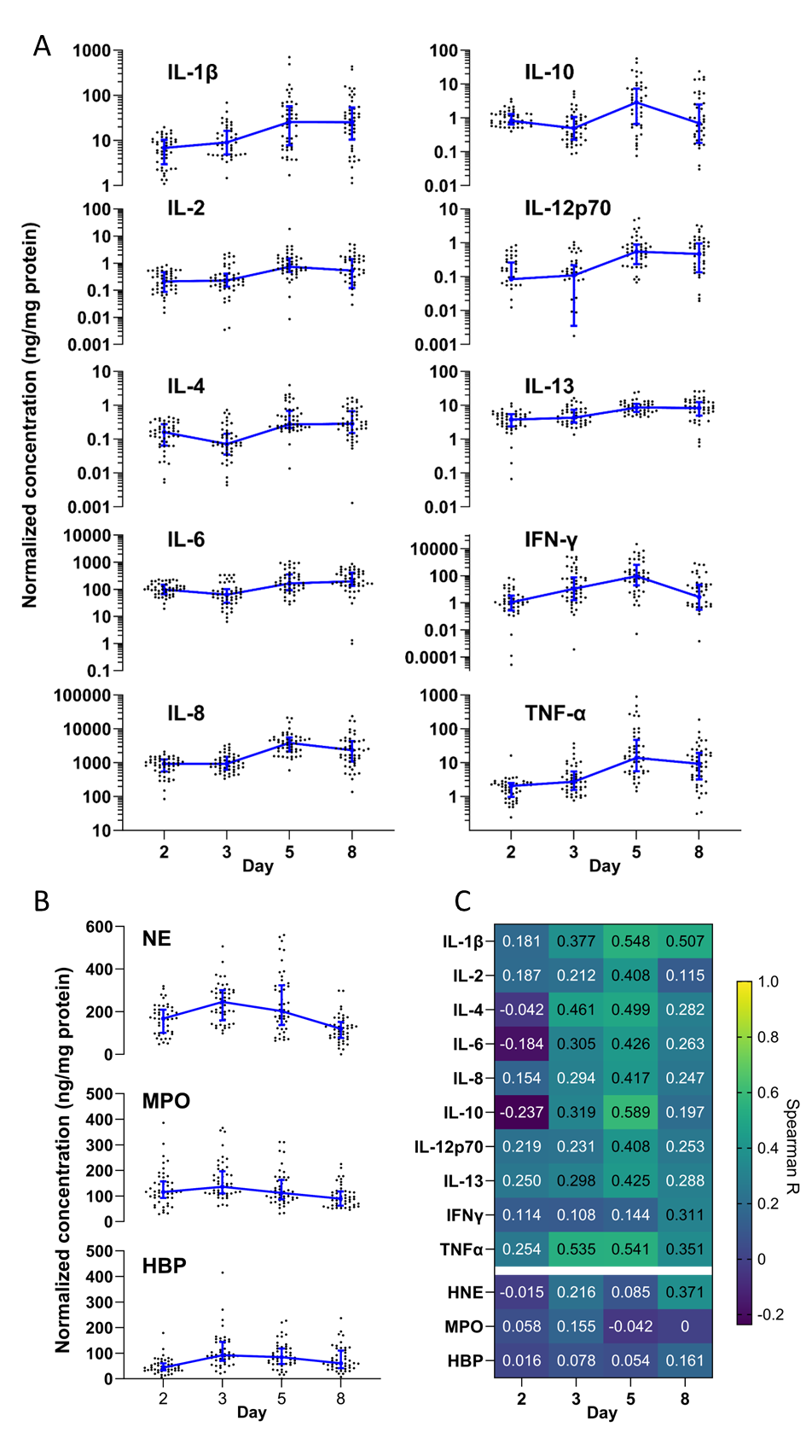
Supplementary Figure 3. **A)** Levels of each cytokine measured in dressing fluid over time normalized for protein content. Each data point is the wound fluid cytokine concentration in a single wound, while the blue line is the median of all wounds that day (n=48), and whiskers are the interquartile range. **B)** Levels of three neutrophil proteins measured in dressing fluid over time normalized for protein content. Each data point is the wound fluid concentration in a single wound, while the blue line is the median of all wounds that day (n=48), and whiskers are the interquartile range. **C)** Spearman correlation coefficients comparing normalized (for protein content) levels of each cytokine and neutrophil protein with bacterial counts in swabs.

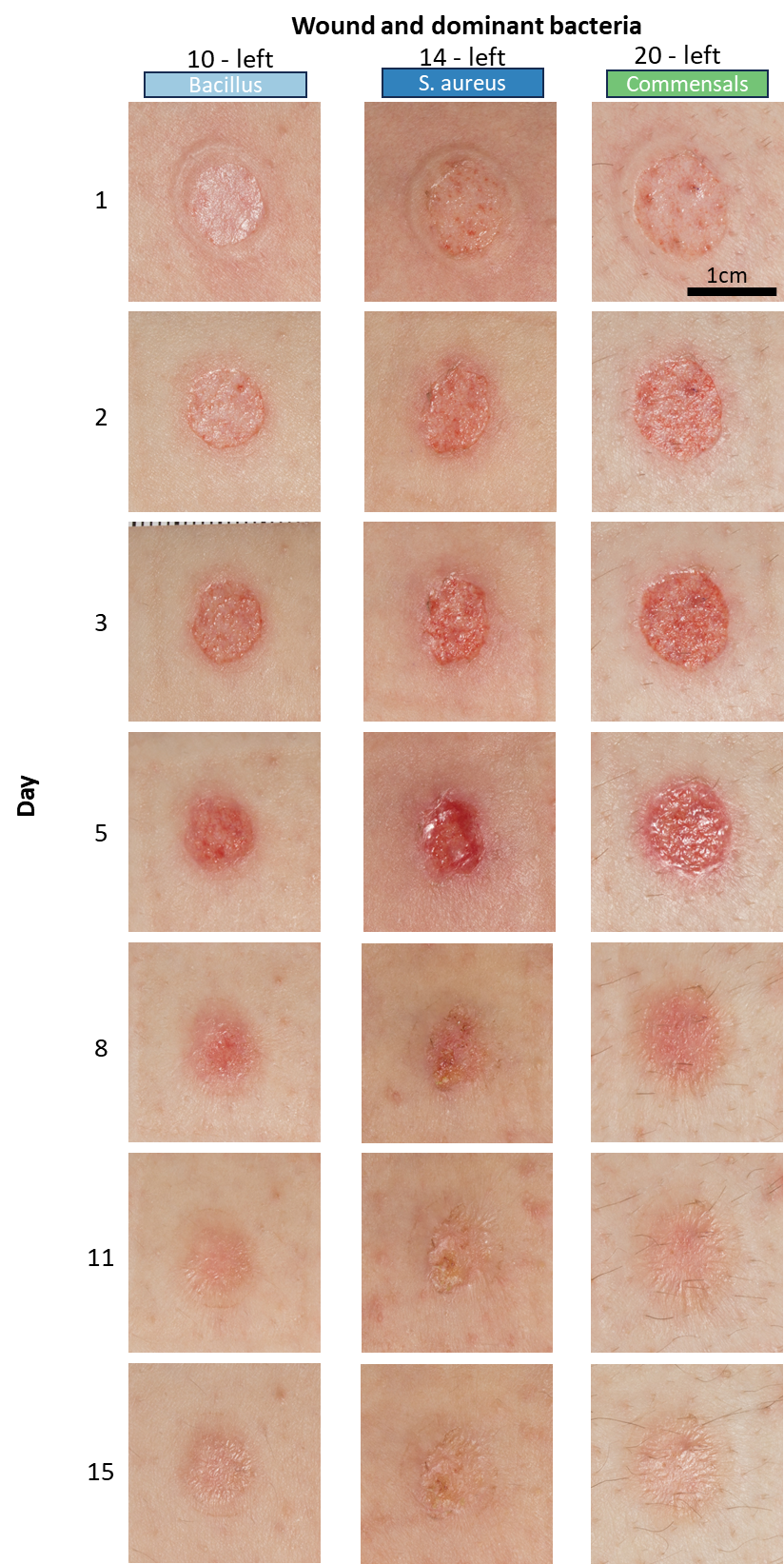

Supplementary Figure 4. Images of three wounds taken over time that had different dominant bacterial strains detected by MALDI-TOF MS (left: *Bacillus sp*.; middle: *S. aureus*; right: various commensals).

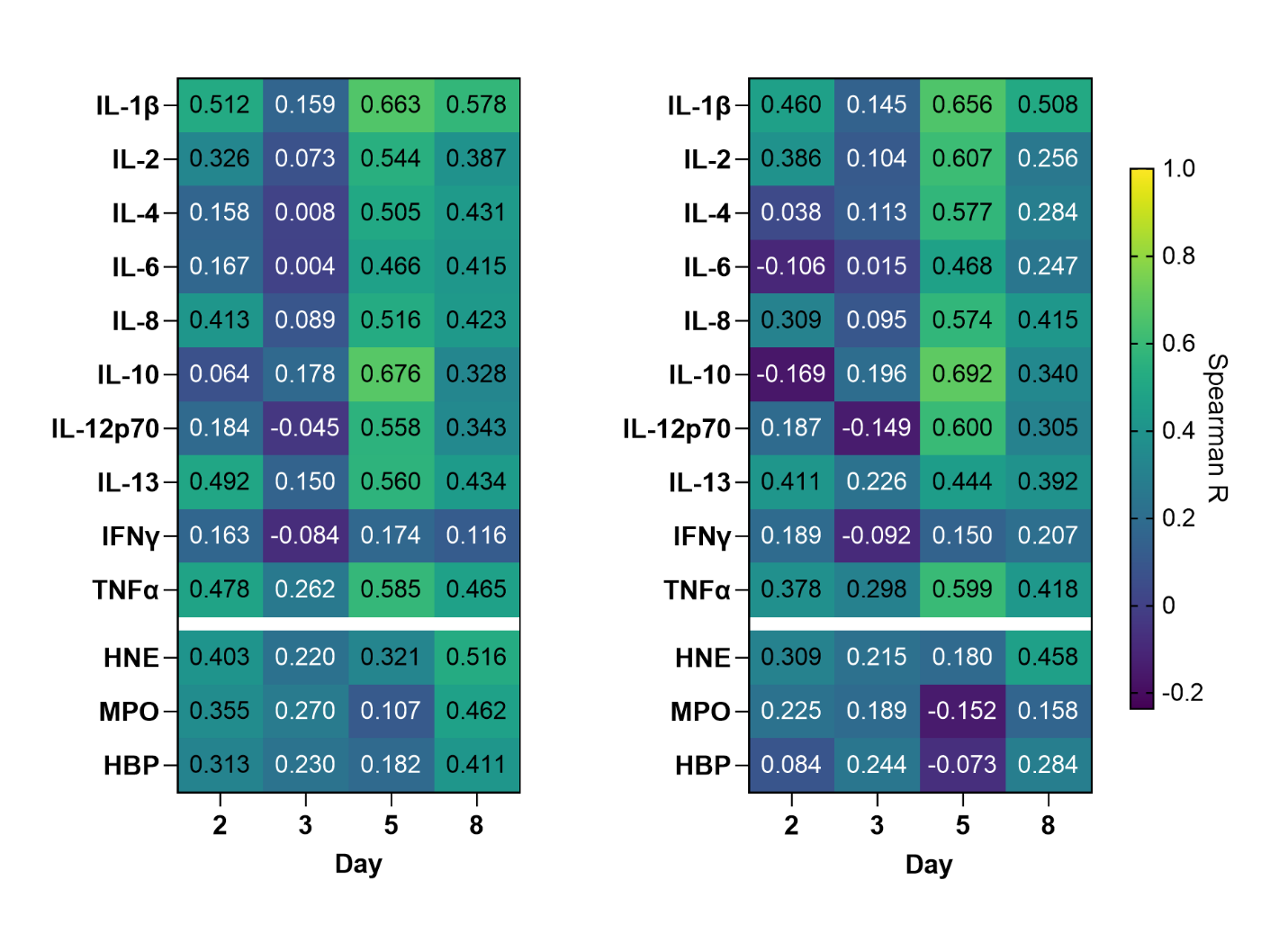

**B**

**A**

**Supplementary Figure 5.** Spearman correlation coefficients comparing normalized (for protein content) levels of each cytokine and neutrophil protein with bacterial counts in dressing fluid.

**Supplementary Figure 5. A)** Spearman correlation coefficients comparing levels of each cytokine and neutrophil protein with quantitative bacterial counts measured in dressing fluid (n=48). **B)** Spearman correlation coefficients comparing normalized (for protein content) levels of each cytokine and neutrophil protein with bacterial counts in dressing fluid.

Supplementary Figure 5. **A)** Spearman correlation coefficients comparing levels of each cytokine and neutrophil protein with quantitative bacterial counts measured in dressing fluid (n=48). **B)** Spearman correlation coefficients comparing normalized (for protein content) levels of each cytokine and neutrophil protein with bacterial counts in dressing fluid.

### Supplementary tables

Supplementary table 1. Median bacterial levels in swab and dressing fluid samples per day.

|  | **Total CFU**  **(median)** | **IQR** | **95% CI** | **CFU/mL**  **(median)** | **CFU/cm^2^**  **(median)** |
| --- | --- | --- | --- | --- | --- |
| *Swab* | | | | | |
| Day 1 | 0 | 0 – 0 | 0 – 0 | 0 | 0 |
| Day 2 | 2871 | 56 – 13208 | 325 – 6667 | 5742 | 7460 |
| Day 3 | 99167 | 9438 – 610417 | 24250 – 349167 | 198334 | 257680 |
| Day 5 | 1054167 | 302083 – 3095834 | 633333 – 1633333 | 2108334 | 2739198 |
| Day 8 | 1683334 | 381250 – 6083334 | 1033333 – 2650000 | 3366667 | 4374055 |
| Day 11 | 1266667 | 445833 – 2787500 | 675000 – 2258333 | 2533333 | 3291367 |
| *Dressing fluid* | | | | | |
| Day 2 | 19312 | 6059 – 61730 | 9357 – 32949 | 8334 | 1042 |
| Day 3 | 1376674 | 314503 – 4283475 | 525549 – 3208660 | 601667 | 75208 |
| Day 5 | 20919514 | 4262095 – 44895112 | 11002813 – 33066067 | 9583333 | 1197917 |
| Day 8 | 15667359 | 5275483 – 33595317 | 7727250 – 24064900 | 7583333 | 947917 |
| Day 11 | 1430953 | 580890 – 3176400 | 854608 – 2172555 | 691667 | 86458 |

CFU = colony forming units, IQR = interquartile range, CI = confidence interval
